# Influence of comorbid diabetes mellitus on outcomes in multiple sclerosis: an English population-based matched cohort study

**DOI:** 10.64898/2026.06.05.26354993

**Authors:** Yolanda Lau, Sedigheh Zabihi, Morghan Hartmann, Georgina Mathlin, Sube Banerjee, Ebaa Marouf, Clare Hadley, Claudia Cooper, Ruth Dobson

## Abstract

**Importance:** As new treatments increase quality and length of life in people with multiple sclerosis (MS), effective prevention and management of common comorbidities, including Diabetes Mellitus (DM), is increasingly important.

**Objective:** To compare incidence of DM and its associations with hospitalisation and mortality in adults with MS and matched controls.

**Design:** Using English primary care data from the Clinical Practice Research Datalink (CPRD), linked to Hospital Episode Statistics and national mortality records, we matched adults with MS diagnosed between 2000 and 2023, with up to ten controls without MS by age, sex, and practice. We excluded individuals with pre-existing DM, defined using diagnostic and management codes. Outcomes included all-cause hospitalisation (number and duration) and mortality. We used Poisson, negative binomial, linear, and Cox proportional hazards models, adjusting for demographic and socioeconomic factors, adding interaction terms to examine if ethnicity, deprivation, and urbanity were associated with outcomes.

**Results:** We included 9,010 individuals with MS and 78,121 matched controls. Over a mean follow-up of 13.2 years, people with MS had over twice the incidence of DM compared with controls (adjusted incidence rate ratio [aIRR]=2.26, 95% CI: 1.96–2.61, *p*<0.001). Among people with MS, incident DM was associated with higher hospitalisation rates (aIRR=1.82, 95%CI: 1.47–2.28, *p*<0.001), longer hospitalisation duration (median 18 vs 4 days, adjusted β=0.53, 95%CI: 0.41–0.65, p<0.001), and increased all-cause mortality when incident DM was modelled as a time-varying exposure (adjusted hazard ratio=1.46, 95%CI: 1.17–1.82, p<0.001), compared to those who did not develop DM. Similar patterns were observed among controls (hospitalisation rates: aIRR = 2.96, 95% CI 2.63–3.23, *p*<0.001; hospitalisation duration: adjusted β = 0.93, 95% CI: 0.86–0.99, *p*<0.001; mortality [time-varying]: HR = 1.50, 95% CI: 1.27–1.77, *p*<0.001). The relationship between DM and increased hospitalisation was stronger in rural areas among those with MS and stronger in White groups among controls.

**Conclusions:** People with MS are more likely to be diagnosed with DM, resulting in greater all-cause hospitalisation and all-cause mortality. This highlights the importance of equitable screening, prevention, and management of DM in people living with MS, with particular attention to geographical health inequalities.

**KEY POINTS:** *Question (one sentence):* Is incident diabetes mellitus (DM) more common in individuals with MS than matched controls and is it associated with all-cause hospitalisation and mortality in this population?

*Findings (1-2 sentence):* In this population-based matched cohort study of 87,131 adults (9,010 with MS), people with MS were twice as likely to be newly diagnosed with DM compared with controls. Among people with MS, a new DM diagnosis was associated with more and longer hospitalisations, and with higher all-cause mortality.

*Meaning (one sentence):* Diabetes prevention, screening, and management has the potential to reduce hospitalisations and extend lives in people living with MS.

## INTRODUCTION

Multiple sclerosis (MS) is a chronic neurological disease with increasing prevalence^1^. Almost 3 million people live with MS globally^2^, of whom 150,000 live in the UK^3^. MS is commonly diagnosed in young adulthood. As life expectancy is only marginally reduced in people living with MS^5^, comorbid physical and mental health conditions may occur. Comorbidities in MS are associated with worse clinical outcomes, including greater and more rapid disability progression^6,7^, reduced access to disease-modifying treatments^6^, delayed diagnosis and treatment of the comorbid conditions themselves^8^ and lower quality of life^9^.

Diabetes mellitus (DM) is a common cardiometabolic condition worldwide, and a major global contributor to morbidity and mortality^10^. Existing evidence on comorbid DM and outcomes in people with MS is limited, predating widespread use of disease-modifying therapies and reporting mixed findings. A 2024 rapid review identified three cohort studies examining the association between DM and mortality in people with MS^6^, with two reporting increased mortality risk among those with DM^11,12^, and one finding no association^13^. A nationwide cohort study in Denmark found no difference in the incidence of DM between those with MS and controls during a median follow-up of 8.3 years^14^. No studies have examined the influence of comorbid incident DM and MS on hospitalisation, an important contributor to decreased quality of life. Focusing on incident disease captures the impact of potential delayed diagnosis and treatment of DM in MS, which may be diluted across prevalent cases. The potential influence of social determinants of health on these outcomes has not been established.

We addressed this evidence gap using English national linked primary and secondary care data. Our objectives were to: (1) compare incidence of DM between people with MS and controls; (2) examine associations between incident DM and all-cause hospitalisation (number and duration); (3) examine associations between incident DM and all-cause mortality; and (4) explore whether health inequalities modify these relationships.

## METHODS

### Study design and ethical approvals

Individuals with MS were matched to controls without MS by sex, year of birth, and practice. This project was approved by the Independent Scientific Advisory Committee for MHRA Database Research (protocol number 24_004057).

### Data source

Participants were identified from the Clinical Practice Research Datalink (CPRD) Gold and Aurum datasets containing anonymised longitudinal primary care data for approximately 60 million patients (∼20% of the UK population) from over 2,400 UK general practices.

We linked CPRD data to Hospital Episode Statistics (HES) (Admitted Patient Care [APC], Outpatient [OP], Accident and Emergency [A&E]), and Office for National Statistics (ONS) death registrations. Area-level characteristics were derived from the 2011 Rural–Urban Classification at Lower Layer Super Output Area (LSOA) level and patient-level Index of Multiple Deprivation (IMD).

### Exposure and cohort definition

The study population was drawn from patients eligible for data linkage across all datasets registered with a primary care practice in England between 2000 and 2023. Exclusion criteria are shown in Table 1.

**Table 1.** Exclusion criteria.

|  |  |
| --- | --- |
| **Year of birth and registration date** | • Absence of a record for a year of birth<br>• An age of greater than 115 at the end of follow-up<br>• An empty or invalid first registration date<br>• A first registration date prior to birth year |
|  | • A transfer out date prior to their first registration date |
| **MS patients** | • Without MS diagnosis date<br>• Without at least two years of follow-up data (defined by patient remained registered with their practice $\geq 2$ years after their MS diagnosis) after the first recorded diagnostic code of MS<br>• A first diagnostic MS code recorded within six months of registration<br>• If an MS patient was excluded, their matched controls were also excluded. |
| **Covariates** | • Missing data on sociodemographic variables (age, sex, Index of Multiple Deprivation [IMD], and rural-urban) |
| **Hospitalisation data** | • Recorded health care episodes in years prior to birth year<br>• Recorded health care episodes with empty or invalid event dates |

#### Exposure

Diagnosis data for MS and DM were identified using multiple data sources, including CPRD and linked secondary data from HES. Code lists for MS and DM were developed using verified clinical code lists^15,16^ and in consultation with clinicians (Table 1 in the Appendix).

##### MS cases

The exposed group included adults aged 18–115 years at the time of an MS code recorded in primary care. MS was defined by the presence of two or more MS diagnostic or management codes, or a single MS code accompanied by an appropriate prescription. Codelists for prescriptions were developed using British National Formulary chapters. The date of MS diagnosis (index date) was defined as the first recorded MS code.

##### Matched controls

Each individual with MS was matched to up to 10 controls without MS. Controls were assigned the same index date as their matched MS case.

##### DM

DM was defined by the presence of at least one diagnostic or management code for type 1, type 2, or unspecified DM, with or without an antidiabetic prescription. Those recorded as having gestational diabetes were excluded. DM was classified as suspected or confirmed: records with a single code, or a prescription without a corresponding diagnosis, were classified as suspected, while diagnoses appearing at least twice or accompanied by an appropriate prescription were confirmed. The date of DM diagnosis was defined as the first recorded diagnostic or management code or prescription. Individuals with DM diagnosed prior to the index date (i.e., prevalent DM) were excluded.

#### Outcomes

A six-month wash-out period after the MS diagnosis was applied to ensure that outcomes were not related to the diagnostic process.

- All-cause hospitalisation episodes and duration: Any hospital admission and the duration of all hospital admissions during the follow-up period were calculated, using HES APC.
- All-cause mortality: Date of death was obtained from ONS death registration records.

#### Covariates

Covariates included age, sex, ethnicity (defined using self-reported UK Census categories), IMD (a measure of socioeconomic status), urban/rural classification (the practice the patient was registered to), and smoking status.

### Missing data

Those with missing age, sex, IMD, or rural/urban classification were excluded, and missing ethnicity data were retained as “unknown”. Data missingness was reported for smoking status. Differences between those with and without missing data were examined using chi-squared tests to assess whether missingness was plausibly consistent with a missing-at-random (MAR) assumption. Significant differences were observed between those with and without smoking status. Smoking status was imputed using multiple imputation by chained equations (MICE), using five imputed datasets.

### Statistical analysis

The analysis plan was pre-registered^17^. Chi-squared tests and t-tests were used to compare baseline characteristics between individuals with MS and their matched controls. Models were adjusted for age, sex, ethnicity, IMD, rural/urbanity, smoking status, and random effects for practices.

#### Objective 1: Incidence of DM

We calculated incidence rates of DM by patient-years. Multivariable Poisson regression models were used to compare the incidence of suspected or confirmed DM diagnosis between groups. Results were reported as incidence rate ratio (IRR).

#### Objective 2: All-cause hospitalisation episodes and duration

We used negative binomial and linear regression models to examine differences in hospitalisation episodes (number and duration respectively) between groups. Separate models were conducted for MS and controls. Results were reported as IRR and coefficients. To examine whether these differed between individuals with MS and controls, an interaction term between MS status and DM was fitted.

#### Objective 3: All-cause mortality

Adjusted Cox proportional hazards regression models were used to compare time to all-cause mortality between individuals with and without DM. Proportional hazards assumption was assessed using Schoenfeld residuals plots and test. Incident DM was modelled as a time-varying exposure. Analyses were conducted separately for MS and controls, and results were reported as hazard ratio (HR). To examine whether hazard ratios differed between individuals with MS and controls, an interaction term between MS and DM was fitted.

#### Objective 4: Potential health inequalities

Interaction terms were added to models to examine if ethnicity, IMD or urbanity modified associations with hospitalisation and mortality. We fitted and tested interactions using the likelihood ratio test. Results were stratified by the factor of interaction (e.g. rural or urban area) where the interaction term was statistically significant.

#### Subgroup and sensitivity analyses

Subgroup analysis was conducted to examine type 1 and type 2 DM. To assess whether the association between MS and incident DM differed by DM type, an interaction term between MS status and DM type was fitted.

Two sensitivity analyses were conducted. First, we restricted the MS group to those with MS diagnosed from 2002 onward, reflecting the implementation of McDonald diagnostic criteria in clinical practice. Second, we defined incident DM based on confirmed DM only, while individuals with suspected DM were reclassified as no DM.

## RESULTS

### Study characteristics

After applying inclusion and exclusion criteria, the final dataset comprised 9,010 participants with MS and 78,121 matched controls (Figure 1). The mean age was 41.6 years (SD = 12.8), and 30.3% (n=26,247) were male (Table 2). Compared to controls, individuals with MS were older, more likely to be from a White ethnic group, and to be a current/ex-smoker. Confirmed and suspected DM were more common in the MS group (n=270 [2.99%] vs. n=742 [0.95%]). Missing data on smoking status were associated with age, ethnicity, IMD, and MS status (all p < 0.001), suggesting data were not missing completely at random.

**Table 2.**
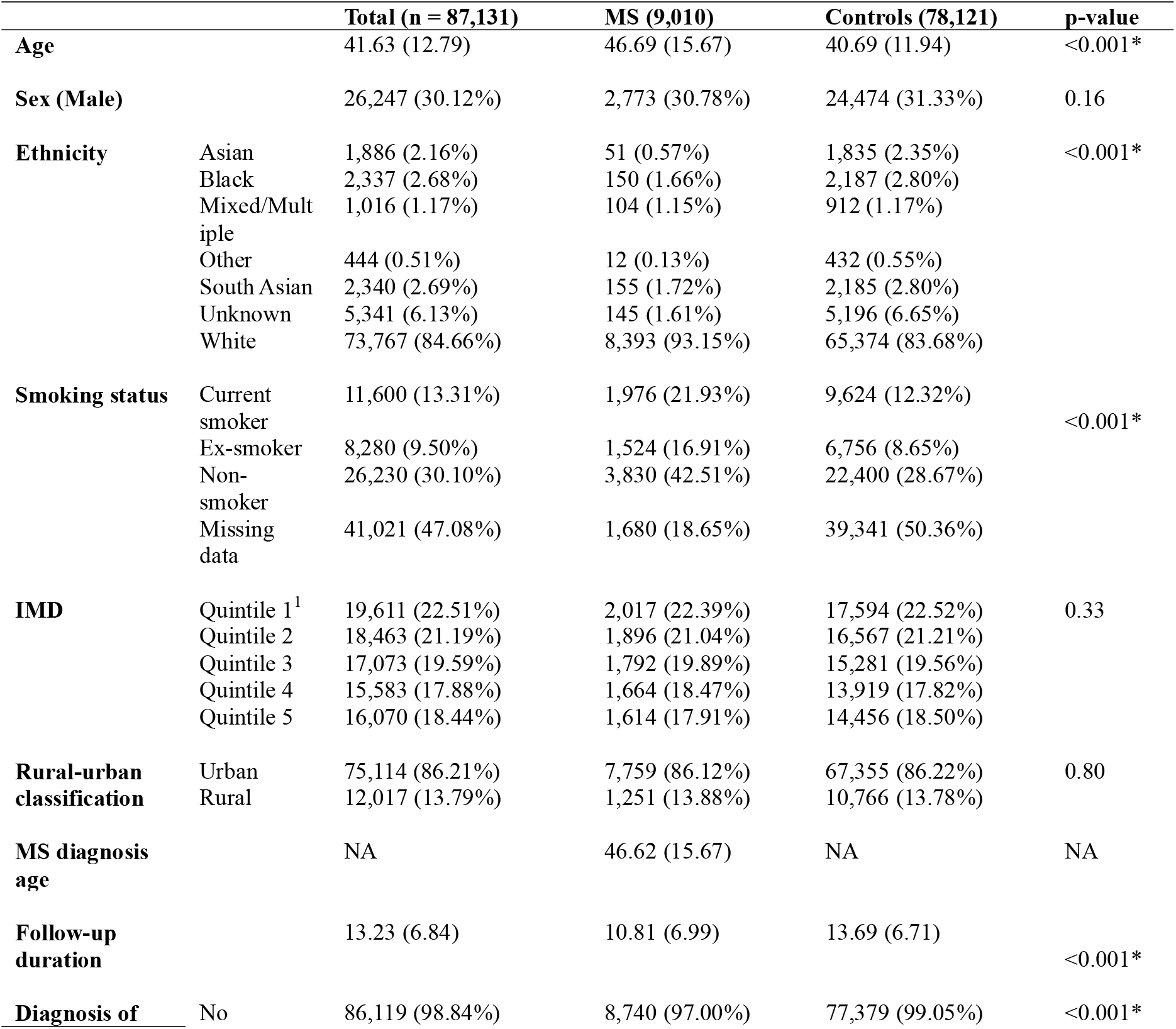
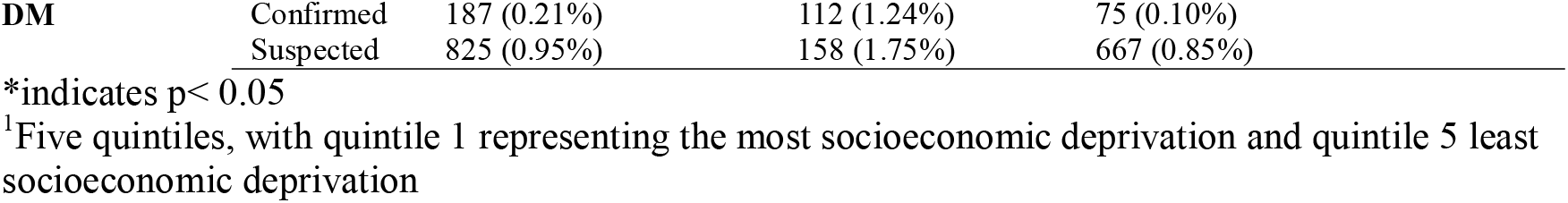
Characteristics of the population at the time of entering the study (index date)

|  |  | Total (n = 87,131) | MS (9,010) | Controls (78,121) | p-value |
| --- | --- | --- | --- | --- | --- |
| **Age** |  | 41.63 (12.79) | 46.69 (15.67) | 40.69 (11.94) | <0.001* |
| **Sex (Male)** |  | 26,247 (30.12%) | 2,773 (30.78%) | 24,474 (31.33%) | 0.16 |
| **Ethnicity** | Asian | 1,886 (2.16%) | 51 (0.57%) | 1,835 (2.35%) | <0.001* |
|  | Black | 2,337 (2.68%) | 150 (1.66%) | 2,187 (2.80%) |  |
|  | Mixed/Multiple | 1,016 (1.17%) | 104 (1.15%) | 912 (1.17%) |  |
|  | Other | 444 (0.51%) | 12 (0.13%) | 432 (0.55%) |  |
|  | South Asian | 2,340 (2.69%) | 155 (1.72%) | 2,185 (2.80%) |  |
|  | Unknown | 5,341 (6.13%) | 145 (1.61%) | 5,196 (6.65%) |  |
|  | White | 73,767 (84.66%) | 8,393 (93.15%) | 65,374 (83.68%) |  |
| **Smoking status** | Current smoker | 11,600 (13.31%) | 1,976 (21.93%) | 9,624 (12.32%) | <0.001* |
|  | Ex-smoker | 8,280 (9.50%) | 1,524 (16.91%) | 6,756 (8.65%) |  |
|  | Non-smoker | 26,230 (30.10%) | 3,830 (42.51%) | 22,400 (28.67%) |  |
|  | Missing data | 41,021 (47.08%) | 1,680 (18.65%) | 39,341 (50.36%) |  |
| **IMD** | Quintile 1<sup>1</sup> | 19,611 (22.51%) | 2,017 (22.39%) | 17,594 (22.52%) | 0.33 |
|  | Quintile 2 | 18,463 (21.19%) | 1,896 (21.04%) | 16,567 (21.21%) |  |
|  | Quintile 3 | 17,073 (19.59%) | 1,792 (19.89%) | 15,281 (19.56%) |  |
|  | Quintile 4 | 15,583 (17.88%) | 1,664 (18.47%) | 13,919 (17.82%) |  |
|  | Quintile 5 | 16,070 (18.44%) | 1,614 (17.91%) | 14,456 (18.50%) |  |
| **Rural-urban classification** | Urban | 75,114 (86.21%) | 7,759 (86.12%) | 67,355 (86.22%) | 0.80 |
|  | Rural | 12,017 (13.79%) | 1,251 (13.88%) | 10,766 (13.78%) |  |
| **MS diagnosis age** |  | NA | 46.62 (15.67) | NA | NA |
| **Follow-up duration** |  | 13.23 (6.84) | 10.81 (6.99) | 13.69 (6.71) | <0.001* |
| **Diagnosis of** | No | 86,119 (98.84%) | 8,740 (97.00%) | 77,379 (99.05%) | <0.001* |

|  |  |  |  |  |
| --- | --- | --- | --- | --- |
| **DM** | Confirmed | 187 (0.21%) | 112 (1.24%) | 75 (0.10%) |
|  | Suspected | 825 (0.95%) | 158 (1.75%) | 667 (0.85%) |

\*indicates $p < 0.05$
<sup>1</sup>Five quintiles, with quintile 1 representing the most socioeconomic deprivation and quintile 5 least socioeconomic deprivation

**Figure 1.**
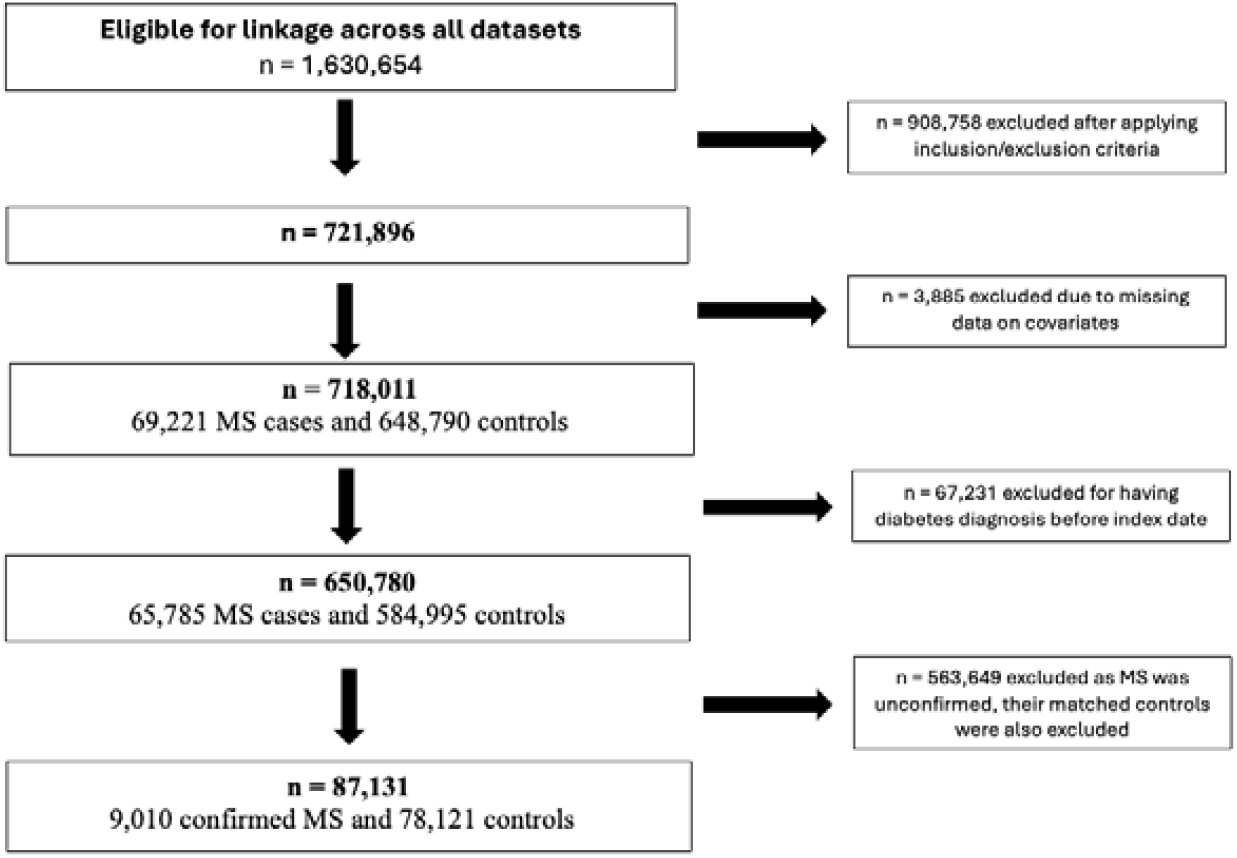
Study flow chart

### Objective 1: Incidence of DM

Over a mean follow-up of 13.2 years, 270 (3.0%) participants with MS and 742 (0.95%) of controls developed DM (suspected or confirmed DM). The crude incidence rate of DM was 2.77 per 1,000 person-years among participants with MS, and 1.12 per 1,000 person-years among controls.

People with MS had around twice the incidence of DM compared with controls (unadjusted IRR = 2.47, 95% CI: 2.15–2.83, p < 0.001; adjusted IRR = 2.26, 95% CI: 1.96–2.61, p < 0.001). Results were consistent in sensitivity analyses (Appendix Table 2).

Among 1,012 participants with incident DM, 749 (74.0%) had information on the type of DM (Type 1: n = 67, Type 2: n = 682). In subgroup analyses by DM known type, this association was not significant (Appendix Table 3).

### Objective 2: All-cause hospitalisation episodes and duration

#### Hospitalisation episodes

Among people with MS, those with incident DM had more hospitalisation episodes in unadjusted (IRR = 1.69, 95% CI: 1.35–2.10, p < 0.001) and adjusted models (adjusted IRR [aIRR] = 1.82, 95% CI: 1.47–2.28, p < 0.001). Results remained significant in sensitivity analyses restricting to MS diagnosed from 2002 onward (Appendix Table 4A). However, in sensitivity analyses using confirmed only, there was no association (Appendix Table 4A).

Among controls, those with incident DM had more hospitalisation episodes in unadjusted (IRR = 3.32, 95% CI 2.97–3.76, p < 0.001) and adjusted models (aIRR = 2.96, 95% CI 2.63–3.23, p < 0.001).

The association between incident DM and hospitalisation episodes was stronger in controls than the MS group (aIRR = 0.47, 95% CI 0.37 – 0.59; p < 0.001).

In both MS group and controls, subgroup analyses found that type 1 and type 2 DM were associated with more hospitalisation episodes (Appendix Table 5A) but the difference between groups was not significant (controls: β = −0.14, 95% CI −0.40 to 0.11; p=0.27; MS: β = −0.07, 95% CI −0.72 to 0.57; p=0.82).

#### Hospitalisation duration

Among people with MS, median duration was 18 days (interquartile range [IQR] 2–87) for those with incident DM and 4 days (IQR 0–22) for those without. This was a significant difference in both unadjusted (β = 0.58, 95% CI: 0.46–0.70, p < 0.001) and adjusted models (β = 0.53, 95% CI: 0.41– 0.65, p < 0.001). Results remained significant in sensitivity analyses on MS diagnosed from 2002 onward (Appendix Table 4B). However, using confirmed DM only, there was no association (Appendix Table 4B).

Among controls, median duration was 10 days (IQR 2–32) for those with incident DM and 1 day (IQR 0–6) for those without incident DM; this was a significant difference in both unadjusted (β = 1.07, 95% CI: 1.00 – 1.14, p < 0.001) and adjusted models (β = 0.93, 95% CI: 0.86 – 0.99, p < 0.001).

The association between incident DM and hospitalisation duration was stronger in the control than the MS group (adjusted β: -0.23 = 95% CI -0.36 – -0.09, p < 0.001).

In both MS group and controls, subgroup analyses found both type 1 and type 2 DM were associated with longer hospitalisation duration (Appendix Table 5B), with no significant difference between groups (controls β = −0.14, 95% CI −0.40 to 0.11, p=0.27; MS: β = −0.07, 95% CI −0.72 to 0.57, p=0.82).

### Objective 3: All-cause mortality

Of the 9,010 participants with MS, 1,582 (17.55%) died during follow-up (94 [5.94%] occurring in the incident DM group). Of the 78,121 controls, 2,618 (3.35%) died (103 [3.93%] occurred in the incident DM group).

Schoenfeld residual tests indicated non-proportional hazards for age (p < 0.001), ethnicity (p = 0.02), and the overall model (p < 0.001) (Figure 1 in Appendix).

Among people with MS, incident DM was not associated with increased all-cause mortality (adjusted HR = 1.21, 95% CI: 0.97 – 1.50, p = 0.10). However, when restricted to MS diagnosed from 2002 onwards, incident DM was associated with a 30% higher hazard of all-cause mortality than those without DM (aHR = 1.30, 95% CI 1.01 – 1.67, p < 0.001). When incident DM was modelled as a time-varying exposure, incident DM was associated with a 46% higher hazard of all-cause mortality (HR = 1.46, 95% CI: 1.17–1.82, p < 0.001). In sensitivity analyses defining incident DM based on confirmed diagnoses, incident DM was associated with higher hazard of all-cause mortality (aHR = 1.58, 95% CI 1.08 – 2.31, p = 0.02, time-varying model: aHR = 2.21, 95% CI 1.40 – 3.48, p < 0.001).

Among controls, incident DM was not associated with mortality relative to those without (HR = 1.15, 95% CI: 0.94–1.41, p = 0.17). When modelled as a time-varying exposure, incident DM was associated with a 49% higher hazard of mortality (HR = 1.50, 95% CI: 1.27–1.77, p < 0.001).

The association between incident DM and mortality did not differ between individuals with MS and controls, either in the main model (HR = 1.06, 95% CI 0.78–1.43, p = 0.71) or when DM was modelled as a time-varying exposure (HR = 0.90, 95% CI 0.68–1.18, p = 0.43).

In the time-varying model, subgroup analyses by DM type showed no significant associations with mortality in MS and controls (Appendix Table 6).

### Objective 4: Findings from interaction analyses

Table 3 shows the results of interaction analyses.

**Table 3.** Results of interaction analyses relating to objectives comparing MS and control groups (p-values)

|  | Ethnicity | IMD | Urban/rural |
| --- | --- | --- | --- |
| **Objective 1: Incidence of DM** | $p = 0.80$ | $p = 0.21$ | $p = 0.71$ |
| **Objective 2: Hospitalisation episodes** | MS: $p = 0.11$<br>Controls: $p = 0.001^*$ | MS: $p = 0.17$<br>Controls: $p = 0.98$ | MS: $p = 0.04^*$<br>Controls: $p = 0.60$ |
| **Objective 2: Hospitalisation duration** | MS: $p = 0.28$<br>Controls: $p = 0.003^*$ | MS: $p = 0.88$<br>Controls: $p = 0.44$ | MS: $p = 0.11$<br>Controls: $p = 0.24$ |
| **Objective 3: Mortality** | MS: $p = 0.66$<br>Controls: $p = 0.52$ | MS: $p = 0.22$<br>Controls: $p = 0.02^*$ | MS: $p = 0.22$<br>Controls: $p = 0.46$ |

Interaction analyses on the adjusted models found no evidence that ethnicity, IMD, or urbanity modified the relationship between incident DM and group (MS or control). In controls, the association between incident DM and hospitalisation episodes and duration was stronger in White than non-White groups (Appendix Tables 7 and 8); while among people with MS, this association was stronger (for number of episodes only) among those living in rural areas (Appendix Table 9). Otherwise, there were no interactions between the associations of incident DM with number of hospitalisation episodes and the sociodemographic factors studied. Among controls from least deprived groups, there was a stronger association between incident DM and mortality (Appendix Table 10).

## DISCUSSION

This study found that people with MS had a higher incidence of DM than matched controls. Among people with MS, incident DM was associated with more hospitalisations and mortality compared with those who did not develop DM. Notably, individuals with MS who developed DM had longer median hospital stay than those without DM. Both people with MS and controls with DM had higher mortality in time-varying models; with no significant difference between these groups. The relationship between DM and greater hospitalisation was stronger in rural areas among those with MS; in controls it was stronger in White ethnic group, and the relationship of DM with mortality was stronger in least deprived groups. Subgroup analyses suggested stronger associations for type 1 DM with incident DM and hospitalisation, however should be interpreted cautiously given small sample sizes.

Our finding that individuals with MS are more likely to develop DM is consistent with research showing they are also at increased risk of cardiovascular disease^18,19^. In both individuals with MS and controls, DM predicted longer hospitalisations, aligning with a study of prevalent DM^20^. A large Canadian study also linked prevalent DM to increased mortality in both people with MS (aHR = 1.47) and matched controls^21^; and two cohort studies reported increased mortality risk among people with MS and prevalent DM, with aHRs of approximately 1.39–1.48^6^.

In controls, there were stronger associations between DM and hospitalisation episodes and duration than in people with MS. One explanation is that people with MS have higher baseline hospitalisation utilisation, for MS-related care^22,23^, so the impact of developing DM on quantity of hospitalisation episodes may be less than for controls. Closer monitoring among people with MS may lead to better management of DM-related complications.

We found evidence that the influence of comorbid DM on outcomes may be modified by health inequalities. Among individuals with MS, the influence of DM on hospitalisation episodes was stronger in those living in rural areas. However, despite evidence that lower socioeconomic status is associated with faster disability progression^24^ and that ethnicity is associated with more symptom burden and overall disability^25^, we found less evidence that IMD or ethnicity modified these associations. More frequent health care contacts among people with MS may attenuate inequalities evident in controls, in whom worse outcomes were associated with deprivation and ethnicity.

### Strengths and limitations

We used a large, population-based matched cohort from linked English primary and secondary care data, enabling robust estimation of DM incidence and its associations with hospitalisation and mortality in MS. Matching on age, sex, and practice reduced confounding, and additional adjustment for socio-demographic factors and smoking further strengthened internal validity. Long follow-up allowed assessment of incident DM and downstream outcomes, and sensitivity analyses—including restriction to incident MS after the McDonald criteria and confirmed diagnoses—supported robustness of findings. Modelling DM as a time-varying exposure reduced immortal time bias.

Limitations should be acknowledged. First, as with all studies using routinely collected electronic health records, the completeness of information may affect generalisability. Ethnicity data are known to be incomplete in routine healthcare records. Second, CPRD relies on healthcare contacts and may under-represent individuals who are less engaged with health services. As a result, people from minority ethnic backgrounds and lower socio-economic groups may be less well represented, which could affect the reliability and precision of some estimates. Finally, subgroup and stratified analyses were based on relatively small sample sizes and should therefore be interpreted with caution.

### Conclusion

Individuals with MS had approximately double the incidence of DM compared with controls. In people with MS, incident DM was associated with higher rates of all-cause hospitalisation and mortality, with similar patterns in controls. The impact of comorbid DM on outcomes in MS varied by health inequalities. These findings highlight the importance of equity-informed DM screening, prevention, and management in people with MS.

## Data Availability

Data were obtained under licence from CPRD

## Acknowledgements

This research is funded through the National Institute for Health and Care Research (NIHR) Policy Research Unit in Dementia and Neurodegeneration – Queen Mary University of London (reference NIHR206110). The views expressed are those of the authors and not necessarily those of the NIHR or the Department of Health and Social Care.

## APPENDIX

**Table 1.**
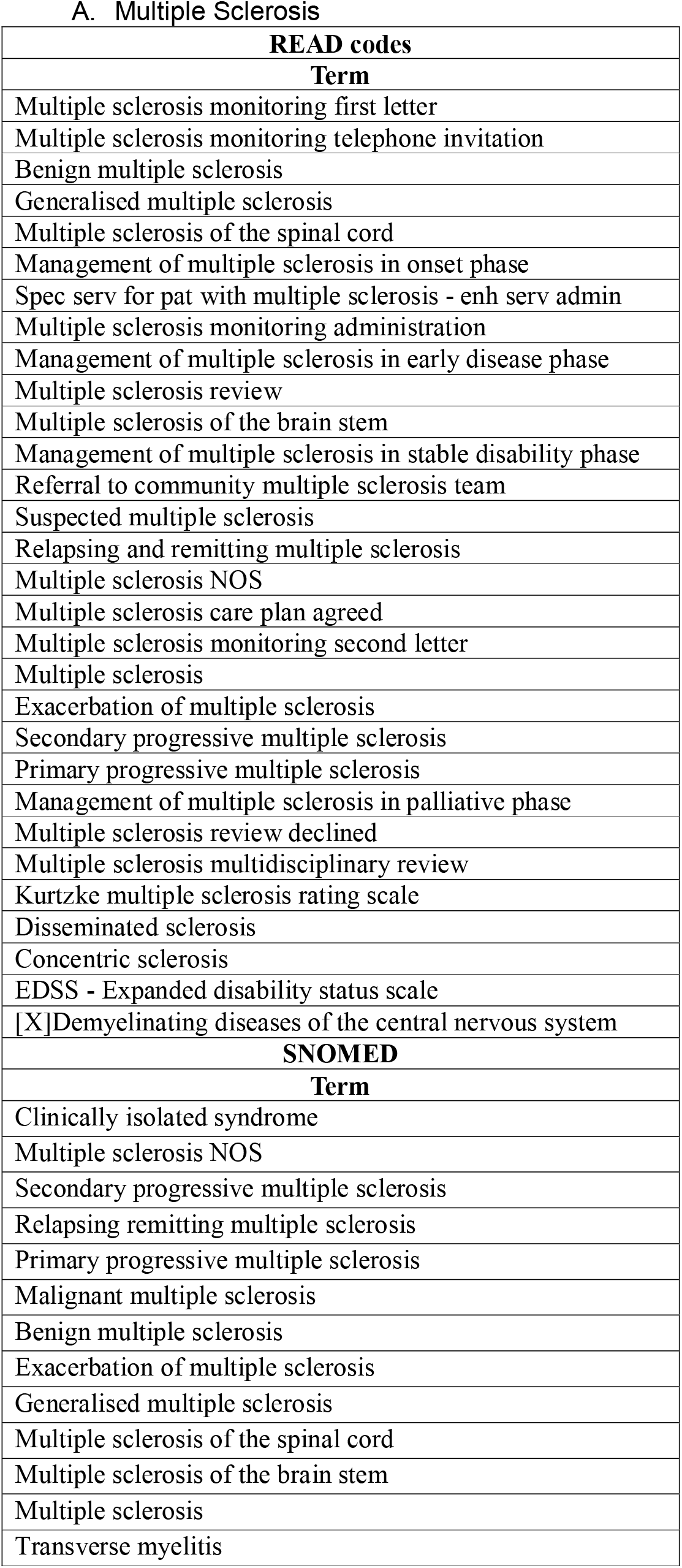
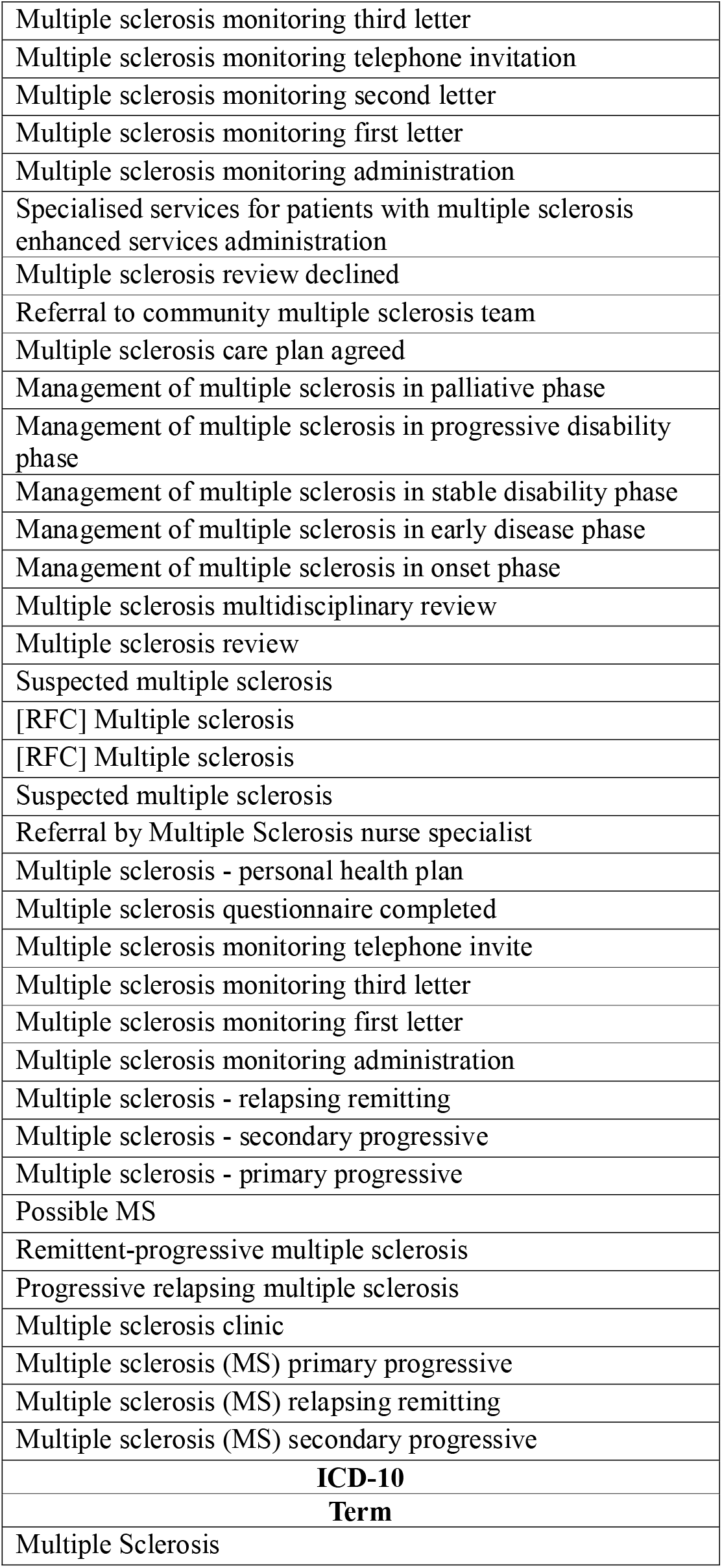
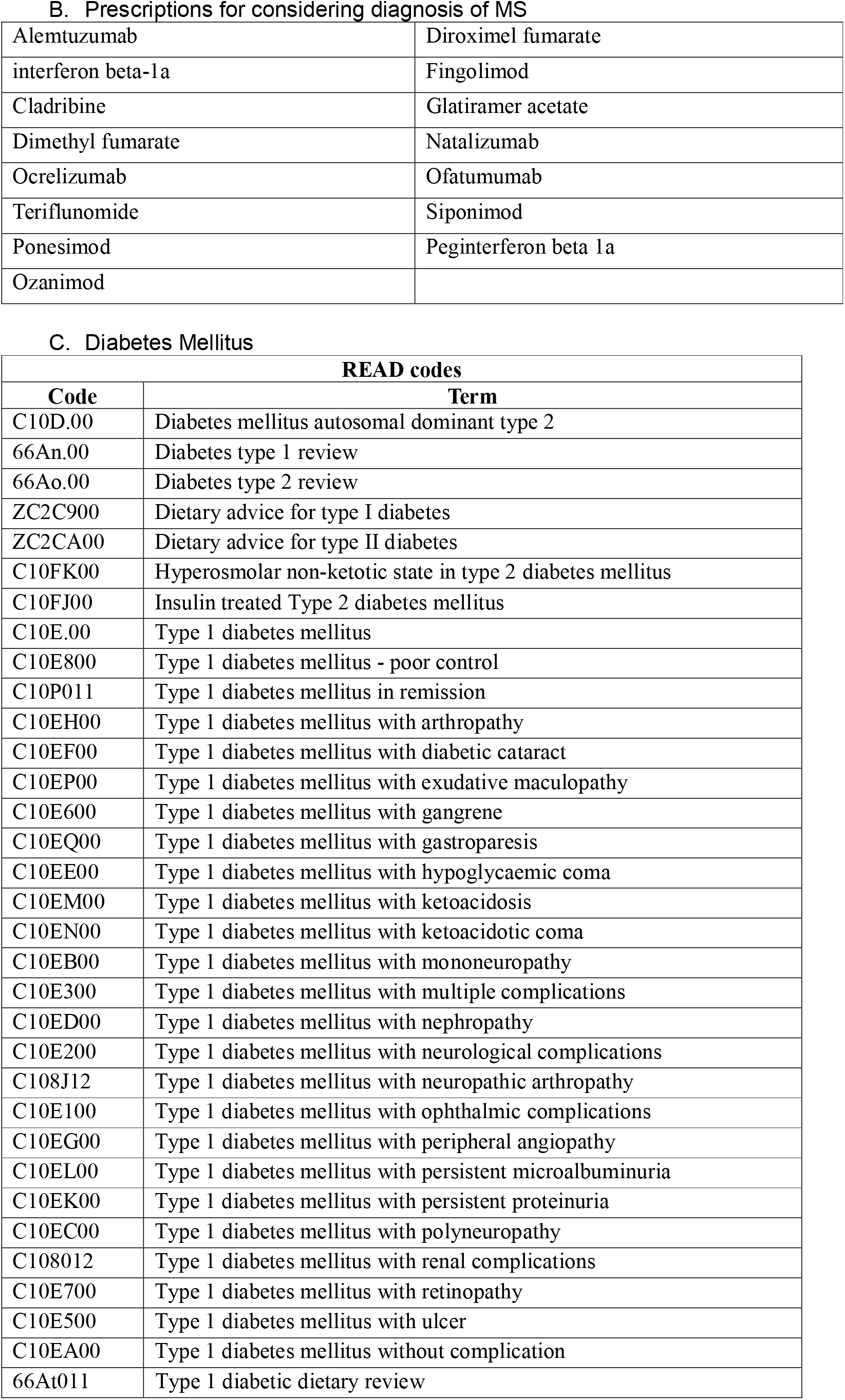
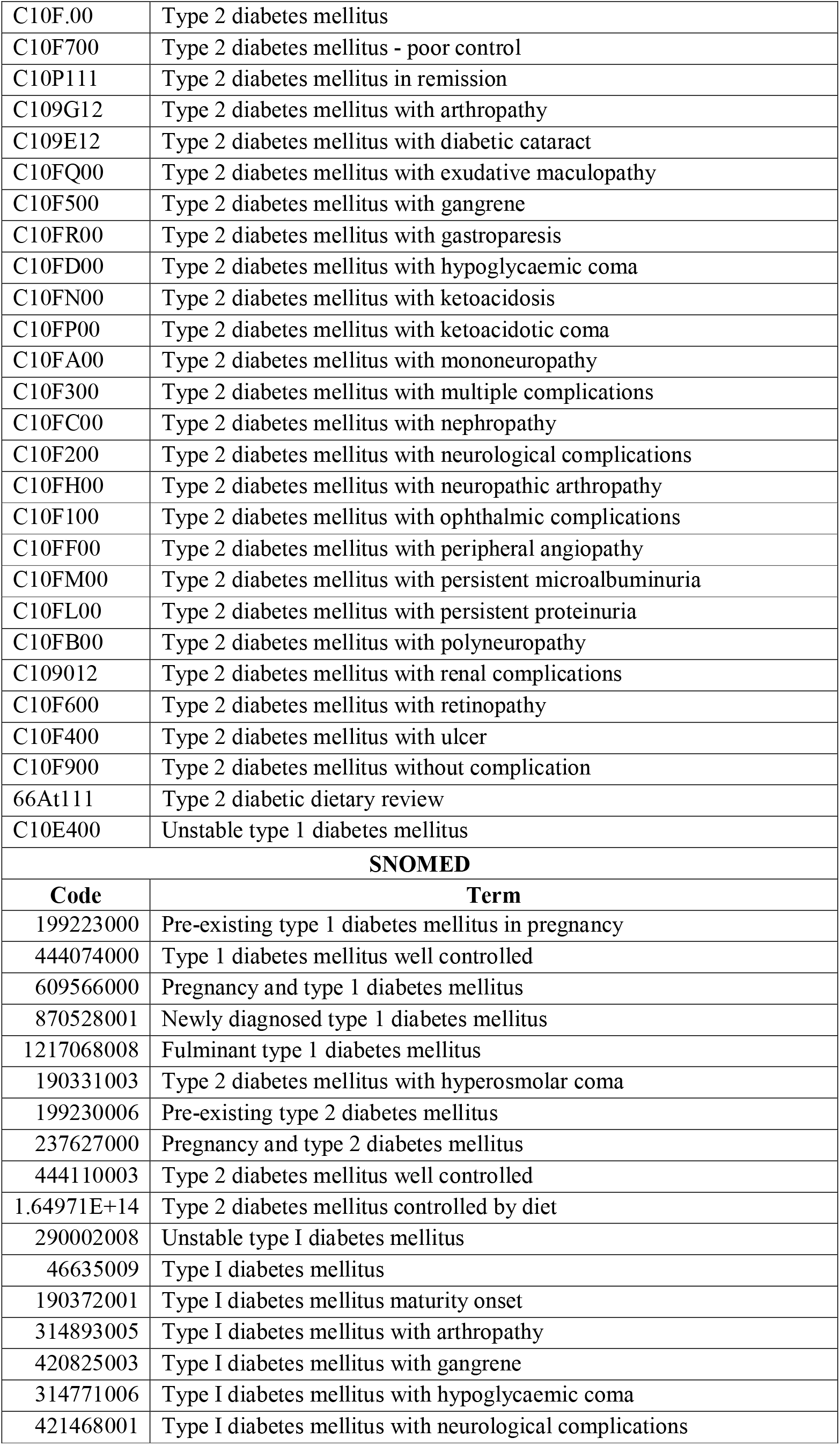
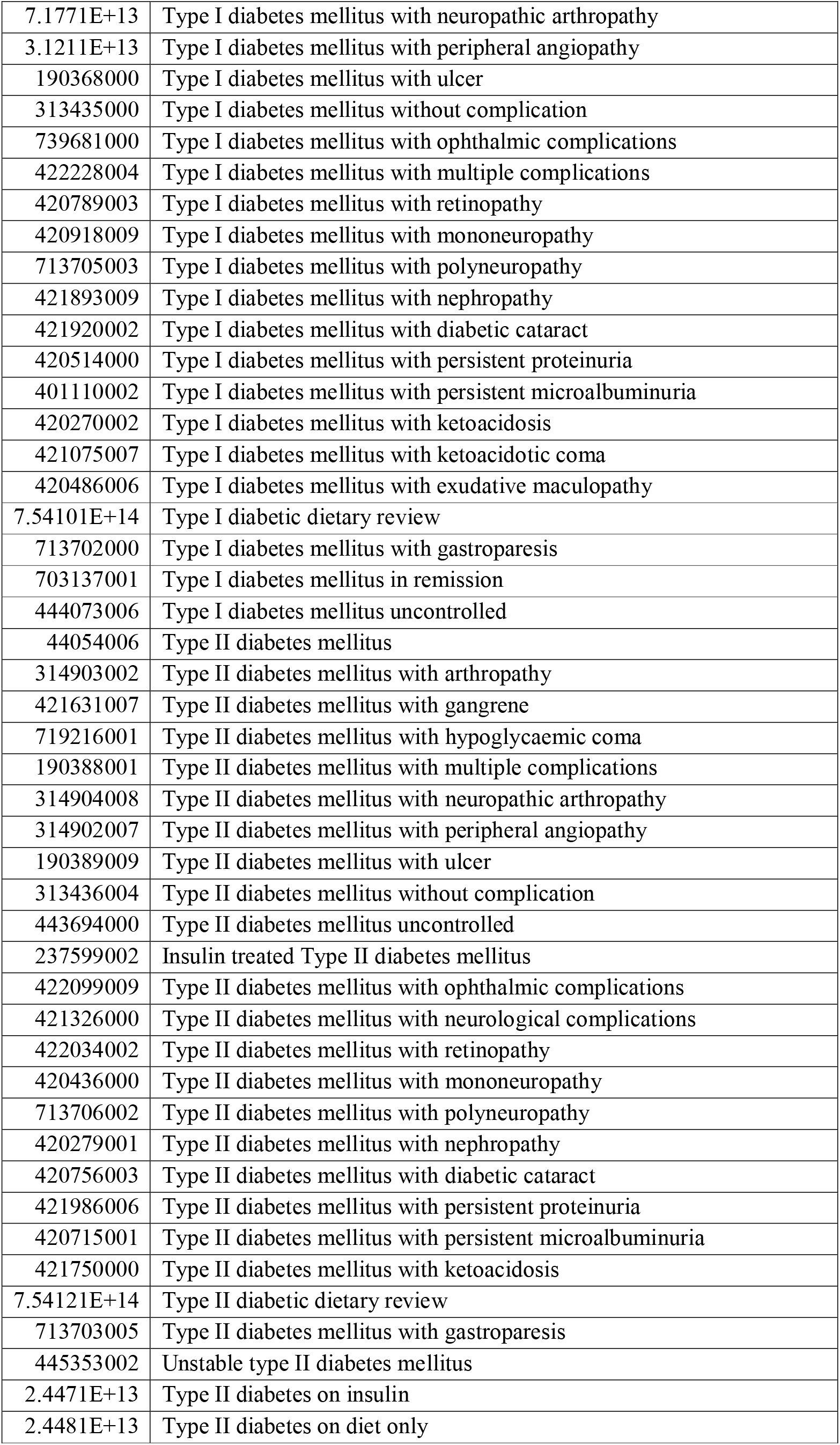
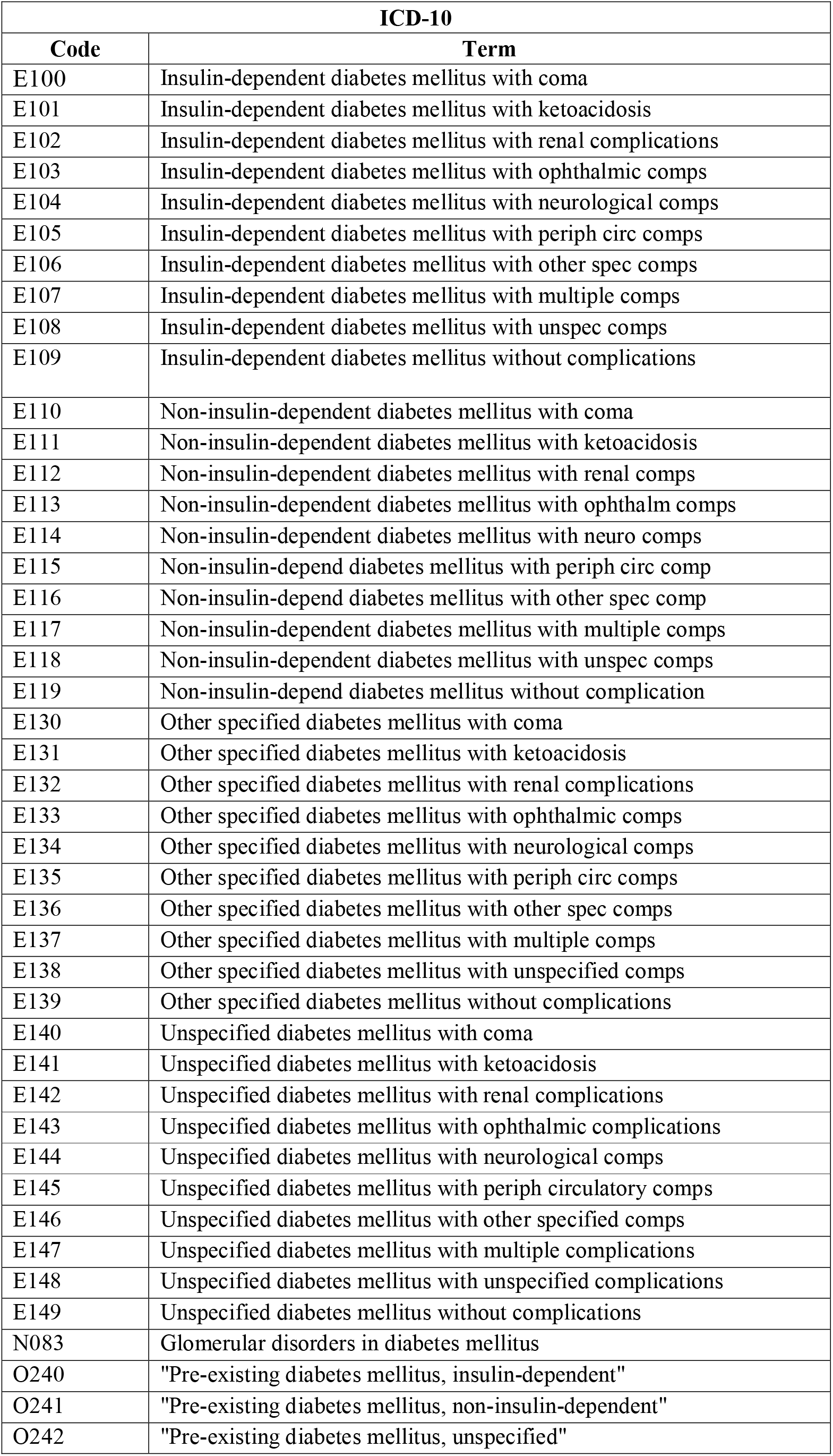
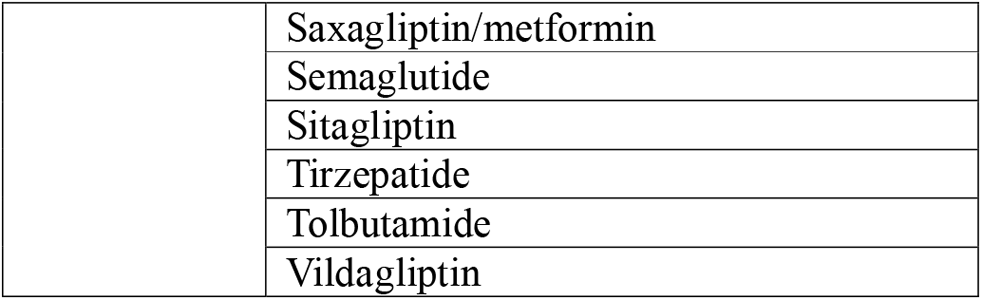
Code lists.

A. Multiple Sclerosis
| READ codes |
| --- |
| Term |
| Multiple sclerosis monitoring first letter |
| Multiple sclerosis monitoring telephone invitation |
| Benign multiple sclerosis |
| Generalised multiple sclerosis |
| Multiple sclerosis of the spinal cord |
| Management of multiple sclerosis in onset phase |
| Spec serv for pat with multiple sclerosis - enh serv admin |
| Multiple sclerosis monitoring administration |
| Management of multiple sclerosis in early disease phase |
| Multiple sclerosis review |
| Multiple sclerosis of the brain stem |
| Management of multiple sclerosis in stable disability phase |
| Referral to community multiple sclerosis team |
| Suspected multiple sclerosis |
| Relapsing and remitting multiple sclerosis |
| Multiple sclerosis NOS |
| Multiple sclerosis care plan agreed |
| Multiple sclerosis monitoring second letter |
| Multiple sclerosis |
| Exacerbation of multiple sclerosis |
| Secondary progressive multiple sclerosis |
| Primary progressive multiple sclerosis |
| Management of multiple sclerosis in palliative phase |
| Multiple sclerosis review declined |
| Multiple sclerosis multidisciplinary review |
| Kurtzke multiple sclerosis rating scale |
| Disseminated sclerosis |
| Concentric sclerosis |
| EDSS - Expanded disability status scale |
| [X]Demyelinating diseases of the central nervous system |
| SNOMED |
| Term |
| Clinically isolated syndrome |
| Multiple sclerosis NOS |
| Secondary progressive multiple sclerosis |
| Relapsing remitting multiple sclerosis |
| Primary progressive multiple sclerosis |
| Malignant multiple sclerosis |
| Benign multiple sclerosis |
| Exacerbation of multiple sclerosis |
| Generalised multiple sclerosis |
| Multiple sclerosis of the spinal cord |
| Multiple sclerosis of the brain stem |
| Multiple sclerosis |
| Transverse myelitis |
| Multiple sclerosis monitoring third letter |
| Multiple sclerosis monitoring telephone invitation |
| Multiple sclerosis monitoring second letter |
| Multiple sclerosis monitoring first letter |
| Multiple sclerosis monitoring administration |
| Specialised services for patients with multiple sclerosis enhanced services administration |
| Multiple sclerosis review declined |
| Referral to community multiple sclerosis team |
| Multiple sclerosis care plan agreed |
| Management of multiple sclerosis in palliative phase |
| Management of multiple sclerosis in progressive disability phase |
| Management of multiple sclerosis in stable disability phase |
| Management of multiple sclerosis in early disease phase |
| Management of multiple sclerosis in onset phase |
| Multiple sclerosis multidisciplinary review |
| Multiple sclerosis review |
| Suspected multiple sclerosis |
| [RFC] Multiple sclerosis |
| [RFC] Multiple sclerosis |
| Suspected multiple sclerosis |
| Referral by Multiple Sclerosis nurse specialist |
| Multiple sclerosis - personal health plan |
| Multiple sclerosis questionnaire completed |
| Multiple sclerosis monitoring telephone invite |
| Multiple sclerosis monitoring third letter |
| Multiple sclerosis monitoring first letter |
| Multiple sclerosis monitoring administration |
| Multiple sclerosis - relapsing remitting |
| Multiple sclerosis - secondary progressive |
| Multiple sclerosis - primary progressive |
| Possible MS |
| Remittent-progressive multiple sclerosis |
| Progressive relapsing multiple sclerosis |
| Multiple sclerosis clinic |
| Multiple sclerosis (MS) primary progressive |
| Multiple sclerosis (MS) relapsing remitting |
| Multiple sclerosis (MS) secondary progressive |
| **ICD-10** |
| **Term** |
| Multiple Sclerosis |

B. Prescriptions for considering diagnosis of MS
|  |  |
| --- | --- |
| Alemtuzumab | Diroximel fumarate |
| interferon beta-1a | Fingolimod |
| Cladribine | Glatiramer acetate |
| Dimethyl fumarate | Natalizumab |
| Ocrelizumab | Ofatumumab |
| Teriflunomide | Siponimod |
| Ponesimod | Peginterferon beta 1a |
| Ozanimod |  |

C. Diabetes Mellitus
| READ codes |  |
| --- | --- |
| Code | Term |
| C10D.00 | Diabetes mellitus autosomal dominant type 2 |
| 66An.00 | Diabetes type 1 review |
| 66Ao.00 | Diabetes type 2 review |
| ZC2C900 | Dietary advice for type I diabetes |
| ZC2CA00 | Dietary advice for type II diabetes |
| C10FK00 | Hyperosmolar non-ketotic state in type 2 diabetes mellitus |
| C10FJ00 | Insulin treated Type 2 diabetes mellitus |
| C10E.00 | Type 1 diabetes mellitus |
| C10E800 | Type 1 diabetes mellitus - poor control |
| C10P011 | Type 1 diabetes mellitus in remission |
| C10EH00 | Type 1 diabetes mellitus with arthropathy |
| C10EF00 | Type 1 diabetes mellitus with diabetic cataract |
| C10EP00 | Type 1 diabetes mellitus with exudative maculopathy |
| C10E600 | Type 1 diabetes mellitus with gangrene |
| C10EQ00 | Type 1 diabetes mellitus with gastroparesis |
| C10EE00 | Type 1 diabetes mellitus with hypoglycaemic coma |
| C10EM00 | Type 1 diabetes mellitus with ketoacidosis |
| C10EN00 | Type 1 diabetes mellitus with ketoacidotic coma |
| C10EB00 | Type 1 diabetes mellitus with mononeuropathy |
| C10E300 | Type 1 diabetes mellitus with multiple complications |
| C10ED00 | Type 1 diabetes mellitus with nephropathy |
| C10E200 | Type 1 diabetes mellitus with neurological complications |
| C108J12 | Type 1 diabetes mellitus with neuropathic arthropathy |
| C10E100 | Type 1 diabetes mellitus with ophthalmic complications |
| C10EG00 | Type 1 diabetes mellitus with peripheral angiopathy |
| C10EL00 | Type 1 diabetes mellitus with persistent microalbuminuria |
| C10EK00 | Type 1 diabetes mellitus with persistent proteinuria |
| C10EC00 | Type 1 diabetes mellitus with polyneuropathy |
| C108012 | Type 1 diabetes mellitus with renal complications |
| C10E700 | Type 1 diabetes mellitus with retinopathy |
| C10E500 | Type 1 diabetes mellitus with ulcer |
| C10EA00 | Type 1 diabetes mellitus without complication |
| 66At011 | Type 1 diabetic dietary review |
| C10F.00 | Type 2 diabetes mellitus |
| C10F700 | Type 2 diabetes mellitus - poor control |
| C10P111 | Type 2 diabetes mellitus in remission |
| C109G12 | Type 2 diabetes mellitus with arthropathy |
| C109E12 | Type 2 diabetes mellitus with diabetic cataract |
| C10FQ00 | Type 2 diabetes mellitus with exudative maculopathy |
| C10F500 | Type 2 diabetes mellitus with gangrene |
| C10FR00 | Type 2 diabetes mellitus with gastroparesis |
| C10FD00 | Type 2 diabetes mellitus with hypoglycaemic coma |
| C10FN00 | Type 2 diabetes mellitus with ketoacidosis |
| C10FP00 | Type 2 diabetes mellitus with ketoacidotic coma |
| C10FA00 | Type 2 diabetes mellitus with mononeuropathy |
| C10F300 | Type 2 diabetes mellitus with multiple complications |
| C10FC00 | Type 2 diabetes mellitus with nephropathy |
| C10F200 | Type 2 diabetes mellitus with neurological complications |
| C10FH00 | Type 2 diabetes mellitus with neuropathic arthropathy |
| C10F100 | Type 2 diabetes mellitus with ophthalmic complications |
| C10FF00 | Type 2 diabetes mellitus with peripheral angiopathy |
| C10FM00 | Type 2 diabetes mellitus with persistent microalbuminuria |
| C10FL00 | Type 2 diabetes mellitus with persistent proteinuria |
| C10FB00 | Type 2 diabetes mellitus with polyneuropathy |
| C109012 | Type 2 diabetes mellitus with renal complications |
| C10F600 | Type 2 diabetes mellitus with retinopathy |
| C10F400 | Type 2 diabetes mellitus with ulcer |
| C10F900 | Type 2 diabetes mellitus without complication |
| 66At111 | Type 2 diabetic dietary review |
| C10E400 | Unstable type 1 diabetes mellitus |
| **SNOMED** |  |
| **Code** | **Term** |
| 199223000 | Pre-existing type 1 diabetes mellitus in pregnancy |
| 444074000 | Type 1 diabetes mellitus well controlled |
| 609566000 | Pregnancy and type 1 diabetes mellitus |
| 870528001 | Newly diagnosed type 1 diabetes mellitus |
| 1217068008 | Fulminant type 1 diabetes mellitus |
| 190331003 | Type 2 diabetes mellitus with hyperosmolar coma |
| 199230006 | Pre-existing type 2 diabetes mellitus |
| 237627000 | Pregnancy and type 2 diabetes mellitus |
| 444110003 | Type 2 diabetes mellitus well controlled |
| 1.64971E+14 | Type 2 diabetes mellitus controlled by diet |
| 290002008 | Unstable type I diabetes mellitus |
| 46635009 | Type I diabetes mellitus |
| 190372001 | Type I diabetes mellitus maturity onset |
| 314893005 | Type I diabetes mellitus with arthropathy |
| 420825003 | Type I diabetes mellitus with gangrene |
| 314771006 | Type I diabetes mellitus with hypoglycaemic coma |
| 421468001 | Type I diabetes mellitus with neurological complications |
| 7.1771E+13 | Type I diabetes mellitus with neuropathic arthropathy |
| 3.1211E+13 | Type I diabetes mellitus with peripheral angiopathy |
| 190368000 | Type I diabetes mellitus with ulcer |
| 313435000 | Type I diabetes mellitus without complication |
| 739681000 | Type I diabetes mellitus with ophthalmic complications |
| 422228004 | Type I diabetes mellitus with multiple complications |
| 420789003 | Type I diabetes mellitus with retinopathy |
| 420918009 | Type I diabetes mellitus with mononeuropathy |
| 713705003 | Type I diabetes mellitus with polyneuropathy |
| 421893009 | Type I diabetes mellitus with nephropathy |
| 421920002 | Type I diabetes mellitus with diabetic cataract |
| 420514000 | Type I diabetes mellitus with persistent proteinuria |
| 401110002 | Type I diabetes mellitus with persistent microalbuminuria |
| 420270002 | Type I diabetes mellitus with ketoacidosis |
| 421075007 | Type I diabetes mellitus with ketoacidotic coma |
| 420486006 | Type I diabetes mellitus with exudative maculopathy |
| 7.54101E+14 | Type I diabetic dietary review |
| 713702000 | Type I diabetes mellitus with gastroparesis |
| 703137001 | Type I diabetes mellitus in remission |
| 444073006 | Type I diabetes mellitus uncontrolled |
| 44054006 | Type II diabetes mellitus |
| 314903002 | Type II diabetes mellitus with arthropathy |
| 421631007 | Type II diabetes mellitus with gangrene |
| 719216001 | Type II diabetes mellitus with hypoglycaemic coma |
| 190388001 | Type II diabetes mellitus with multiple complications |
| 314904008 | Type II diabetes mellitus with neuropathic arthropathy |
| 314902007 | Type II diabetes mellitus with peripheral angiopathy |
| 190389009 | Type II diabetes mellitus with ulcer |
| 313436004 | Type II diabetes mellitus without complication |
| 443694000 | Type II diabetes mellitus uncontrolled |
| 237599002 | Insulin treated Type II diabetes mellitus |
| 422099009 | Type II diabetes mellitus with ophthalmic complications |
| 421326000 | Type II diabetes mellitus with neurological complications |
| 422034002 | Type II diabetes mellitus with retinopathy |
| 420436000 | Type II diabetes mellitus with mononeuropathy |
| 713706002 | Type II diabetes mellitus with polyneuropathy |
| 420279001 | Type II diabetes mellitus with nephropathy |
| 420756003 | Type II diabetes mellitus with diabetic cataract |
| 421986006 | Type II diabetes mellitus with persistent proteinuria |
| 420715001 | Type II diabetes mellitus with persistent microalbuminuria |
| 421750000 | Type II diabetes mellitus with ketoacidosis |
| 7.54121E+14 | Type II diabetic dietary review |
| 713703005 | Type II diabetes mellitus with gastroparesis |
| 445353002 | Unstable type II diabetes mellitus |
| 2.4471E+13 | Type II diabetes on insulin |
| 2.4481E+13 | Type II diabetes on diet only |

| ICD-10 |  |
| --- | --- |
| Code | Term |
| E100 | Insulin-dependent diabetes mellitus with coma |
| E101 | Insulin-dependent diabetes mellitus with ketoacidosis |
| E102 | Insulin-dependent diabetes mellitus with renal complications |
| E103 | Insulin-dependent diabetes mellitus with ophthalmic comps |
| E104 | Insulin-dependent diabetes mellitus with neurological comps |
| E105 | Insulin-dependent diabetes mellitus with periph circ comps |
| E106 | Insulin-dependent diabetes mellitus with other spec comps |
| E107 | Insulin-dependent diabetes mellitus with multiple comps |
| E108 | Insulin-dependent diabetes mellitus with unspec comps |
| E109 | Insulin-dependent diabetes mellitus without complications |
| E110 | Non-insulin-dependent diabetes mellitus with coma |
| E111 | Non-insulin-dependent diabetes mellitus with ketoacidosis |
| E112 | Non-insulin-dependent diabetes mellitus with renal comps |
| E113 | Non-insulin-dependent diabetes mellitus with ophthalm comps |
| E114 | Non-insulin-dependent diabetes mellitus with neuro comps |
| E115 | Non-insulin-depend diabetes mellitus with periph circ comp |
| E116 | Non-insulin-depend diabetes mellitus with other spec comp |
| E117 | Non-insulin-dependent diabetes mellitus with multiple comps |
| E118 | Non-insulin-dependent diabetes mellitus with unspec comps |
| E119 | Non-insulin-depend diabetes mellitus without complication |
| E130 | Other specified diabetes mellitus with coma |
| E131 | Other specified diabetes mellitus with ketoacidosis |
| E132 | Other specified diabetes mellitus with renal complications |
| E133 | Other specified diabetes mellitus with ophthalmic comps |
| E134 | Other specified diabetes mellitus with neurological comps |
| E135 | Other specified diabetes mellitus with periph circ comps |
| E136 | Other specified diabetes mellitus with other spec comps |
| E137 | Other specified diabetes mellitus with multiple comps |
| E138 | Other specified diabetes mellitus with unspecified comps |
| E139 | Other specified diabetes mellitus without complications |
| E140 | Unspecified diabetes mellitus with coma |
| E141 | Unspecified diabetes mellitus with ketoacidosis |
| E142 | Unspecified diabetes mellitus with renal complications |
| E143 | Unspecified diabetes mellitus with ophthalmic complications |
| E144 | Unspecified diabetes mellitus with neurological comps |
| E145 | Unspecified diabetes mellitus with periph circulatory comps |
| E146 | Unspecified diabetes mellitus with other specified comps |
| E147 | Unspecified diabetes mellitus with multiple complications |
| E148 | Unspecified diabetes mellitus with unspecified complications |
| E149 | Unspecified diabetes mellitus without complications |
| N083 | Glomerular disorders in diabetes mellitus |
| O240 | "Pre-existing diabetes mellitus, insulin-dependent" |
| O241 | "Pre-existing diabetes mellitus, non-insulin-dependent" |
| O242 | "Pre-existing diabetes mellitus, unspecified" |

D. List of drugs for consider diabetes
|  |  |
| --- | --- |
| Insulin | Biphasic insulin aspart |
|  | Biphasic insulin lispro |
|  | Biphasic isophane insulin |
|  | Insulin aspart |
|  | Insulin degludec |
|  | Insulin detemir |
|  | Insulin glargine |
|  | Insulin glargine/lixisenatide |
|  | Insulin glulisine |
|  | Insulin human |
|  | Insulin Lispro |
|  | Insulin zinc suspension |
|  | Isophane insulin |
|  | Protamine zinc insulin |
|  | Soluble insulin (Neutral insulin) |
| Antidiabetic | Acarbose |
|  | Albiglutide |
|  | Alogliptin |
|  | Alogliptin/metformin |
|  | Canagliflozin |
|  | Canagliflozin/metformin |
|  | Chlorpropamide |
|  | Dapagliflozin |
|  | Dapagliflozin/metformin |
|  | Dulaglutide |
|  | Empagliflozin |
|  | Empagliflozin/linagliptin |
|  | Empagliflozin/metformin |
|  | Ertugliflozin |
|  | Exenatide |
|  | Glibenclamide |
|  | Gliclazide |
|  | Glimepiride |
|  | Glipizide |
|  | Guar gum |
|  | Ins degludec/liraglutide |
|  | Linagliptin |
|  | Linagliptin/metformin |
|  | Liraglutide |
|  | Lixisenatide |
|  | Metformin hydrochloride |
|  | Metformin hydrochloride/pioglitazone |
|  | Metformin hydrochloride/rosiglitazone |
|  | Metformin hydrochloride/sitagliptin |
|  | Metformin hydrochloride/vildagliptin |
|  | Nateglinide |
|  | Pioglitazone hydrochloride |
|  | Repaglinide |
|  | Rosiglitazone |
|  | Saxagliptin |
|  | Saxagliptin/dapagliflozin |
|  | Saxagliptin/metformin |
|  | Semaglutide |
|  | Sitagliptin |
|  | Tirzepatide |
|  | Tolbutamide |
|  | Vildagliptin |

**Table 2.** Objective 1: Sensitivity analyses.

|  | IRR (95% CI) | p-value |
| --- | --- | --- |
| **MS diagnosed from 2002 onwards (and their matched controls)** |  |  |
| Total n = 78,544, 8,119 with MS and 70,425 controls |  |  |
| Unadjusted | 3.41 (2.84 – 3.93) | <0.001* |
| Adjusted | 3.10 (2.62 – 3.67) | <0.001* |
| **Confirmed DM** |  |  |
| Confirmed DM: MS n = 112; controls n = 75 |  |  |
| Unadjusted | 10.12 (7.55 – 13.56) | <0.001* |
| Adjusted | 9.06 (6.68 – 12.29) | <0.001* |

**Table 3.**
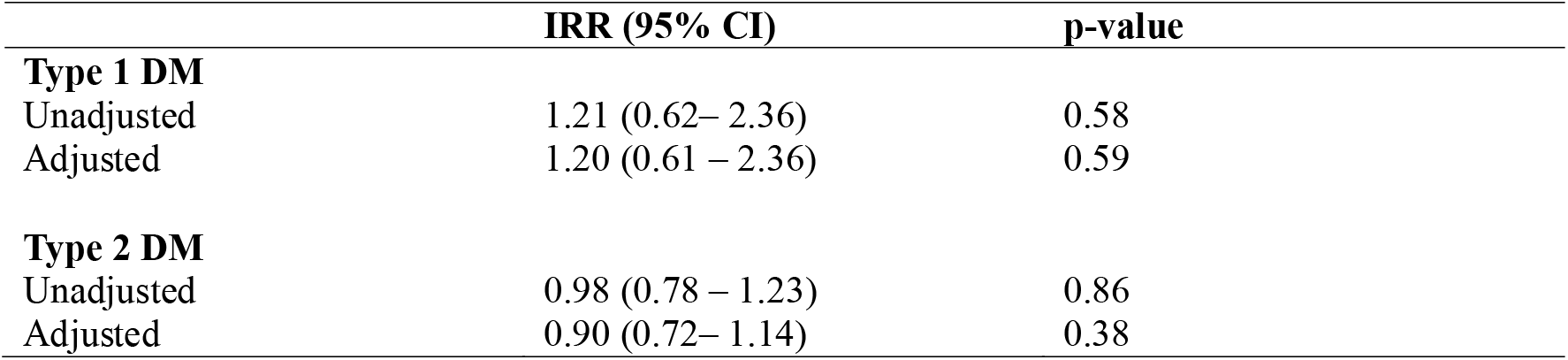
Objective 1: Subgroup analyses.

|  | IRR (95% CI) | p-value |
| --- | --- | --- |
| **Type 1 DM** |  |  |
| Unadjusted | 1.21 (0.62– 2.36) | 0.58 |
| Adjusted | 1.20 (0.61 – 2.36) | 0.59 |
| **Type 2 DM** |  |  |
| Unadjusted | 0.98 (0.78 – 1.23) | 0.86 |
| Adjusted | 0.90 (0.72– 1.14) | 0.38 |

**Table 4.** Objective 2: Sensitivity analyses.

|  | B (95% CI) | p-value |
| --- | --- | --- |
| **MS diagnosed from 2002 onwards** |  |  |
| MS n = 8,119 |  |  |
| Unadjusted | 1.67 (1.30 – 2.15) | <0.001* |
| Adjusted | 1.80 (1.41 – 2.32) | <0.001* |
| **Confirmed DM** |  |  |
| Total n = 9,010, 112 with confirmed DM, 8,898 no DM |  |  |
| Unadjusted | 0.97 (0.69 – 1.37) | 0.87 |
| Adjusted | 1.05 (0.74 – 1.47) | 0.80 |

**B. Hospitalisation duration**
|  | B (95% CI) | p-value |
| --- | --- | --- |
| **MS diagnosed from 2002 onwards** |  |  |
| MS n = 8,119 |  |  |
| Unadjusted | 0.59 (0.46 - 0.73) | <0.001* |
| Adjusted | 0.54 (0.41 – 0.67) | <0.001* |
| **Confirmed DM** |  |  |
| Total n = 9,010, 112 with confirmed DM, 8,898 no DM |  |  |
| Unadjusted | -0.001 (-0.02 – 0.02) | 0.90 |
| Adjusted | -0.14 (-0.44 – 0.17) | 0.03* |

**Table 5.**
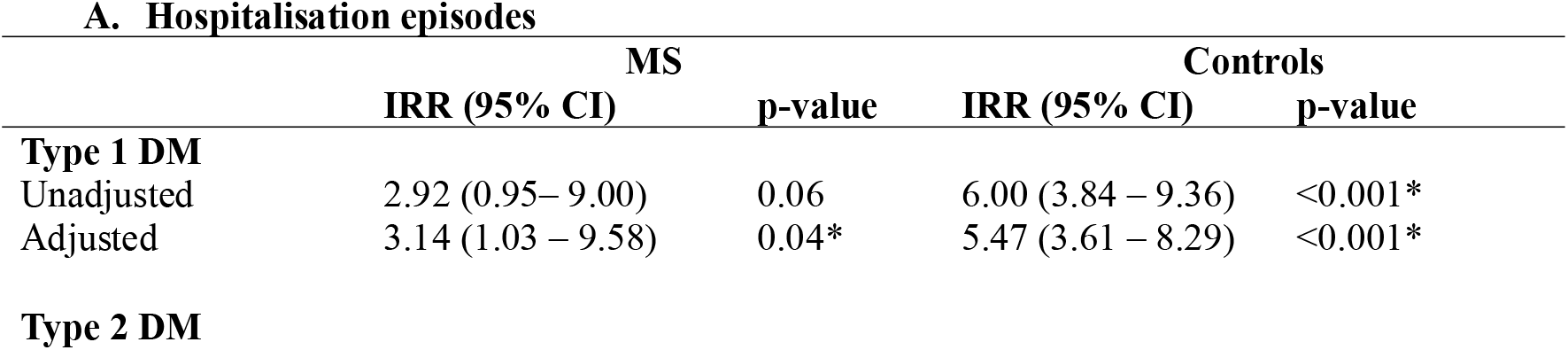
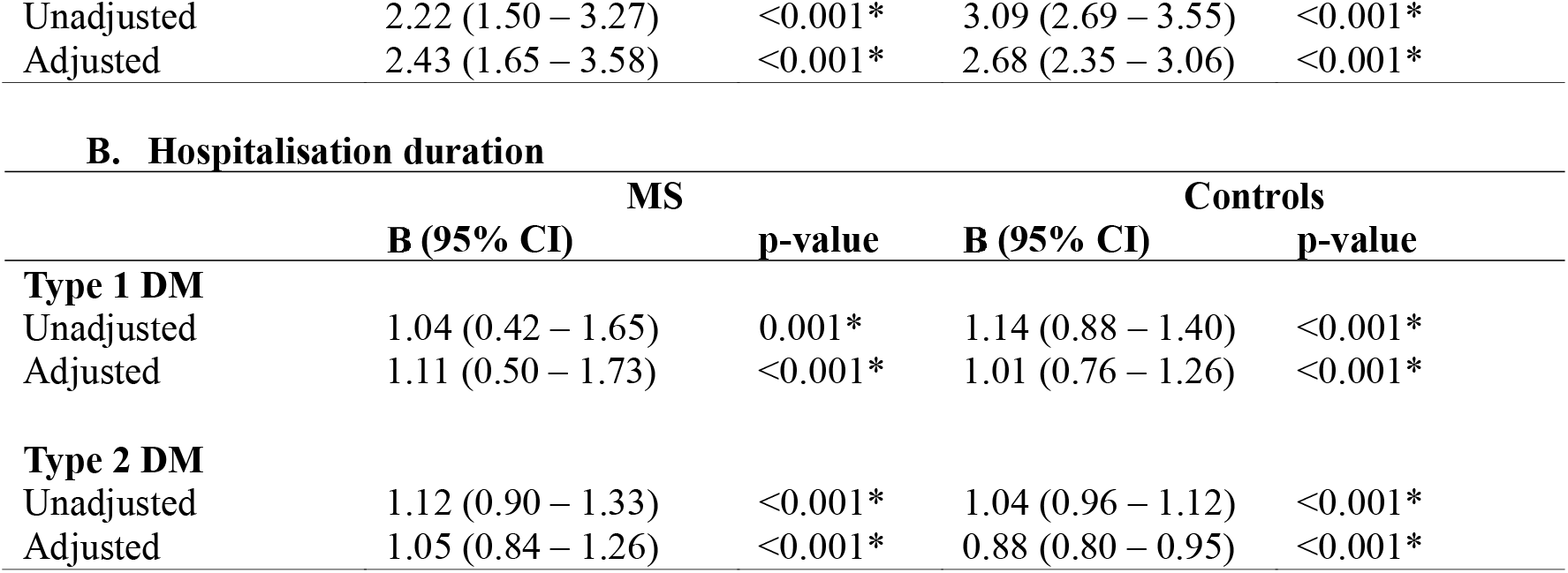
Objective 2: Subgroup analyses.

|  | MS |  | Controls |  |
| --- | --- | --- | --- | --- |
|  | IRR (95% CI) | p-value | IRR (95% CI) | p-value |
| **Type 1 DM** |  |  |  |  |
| Unadjusted | 2.92 (0.95– 9.00) | 0.06 | 6.00 (3.84 – 9.36) | <0.001* |
| Adjusted | 3.14 (1.03 – 9.58) | 0.04* | 5.47 (3.61 – 8.29) | <0.001* |
| **Type 2 DM** |  |  |  |  |
| Unadjusted | 2.22 (1.50 – 3.27) | <0.001* | 3.09 (2.69 – 3.55) | <0.001* |
| Adjusted | 2.43 (1.65 – 3.58) | <0.001* | 2.68 (2.35 – 3.06) | <0.001* |

B. Hospitalisation duration
|  | MS |  | Controls |  |
| --- | --- | --- | --- | --- |
|  | B (95% CI) | p-value | B (95% CI) | p-value |
| **Type 1 DM** |  |  |  |  |
| Unadjusted | 1.04 (0.42 – 1.65) | 0.001* | 1.14 (0.88 – 1.40) | <0.001* |
| Adjusted | 1.11 (0.50 – 1.73) | <0.001* | 1.01 (0.76 – 1.26) | <0.001* |
| **Type 2 DM** |  |  |  |  |
| Unadjusted | 1.12 (0.90 – 1.33) | <0.001* | 1.04 (0.96 – 1.12) | <0.001* |
| Adjusted | 1.05 (0.84 – 1.26) | <0.001* | 0.88 (0.80 – 0.95) | <0.001* |

**Table 6.** Objective 3: Subgroup analyses (time-varying model)

|  | MS |  | Controls |  |
| --- | --- | --- | --- | --- |
|  | HR (95% CI) | p-value | HR (95% CI) | p-value |
| **Type 1 DM** |  |  |  |  |
|  | 1.09 (0.15 – 8.12) | 0.94 | 1.67 (0.88– 3.19) | 0.12 |
| **Type 2 DM** |  |  |  |  |
|  | 0.96 (0.67 – 1.37) | 0.82 | 1.21 (0.96 – 1.61) | 0.10 |

**Table 7.** Objective 2: Stratified analyses: Hospitalisation episodes in control group Stratified by ethnicity.

|  | IRR (95% CI) | p-value |
| --- | --- | --- |
| **White** |  |  |
| Unadjusted | 3.55 (2.93 – 3.82) | <0.001* |
| Adjusted | 2.95 (2.59 – 3.35) | <0.001* |
| **Non-White** |  |  |
| Unadjusted | 1.84 (1.89 – 3.57) | <0.001* |
| Adjusted | 2.28 (1.69 – 3.09) | <0.001* |
| Adjusted for age, gender, IMD, and smoking status |  |  |

**Table 8.** Objective 2 Stratified analyses: Hospitalisation duration in controls Stratified by ethnicity.

|  | B (95% CI) | p-value |
| --- | --- | --- |
| **White** |  |  |
| Unadjusted | 1.09 (1.00 – 0.17) | <0.001* |
| Adjusted | 0.95 (0.87 – 1.03) | <0.001* |
| **Non-White** |  |  |
| Unadjusted | 0.91 (0.75 – 1.07) | <0.001* |
| Adjusted | 0.81 (0.64 – 0.97) | <0.001* |
| Adjusted for age, gender, IMD, and smoking status |  |  |

**Table 9.** Objective 2: Stratified analyses: Hospitalisation episodes in MS group Stratified by urbanity.

|  | IRR (95% CI) | p-value |
| --- | --- | --- |
| **Urban** |  |  |
| Unadjusted | 1.52 (1.20-1.94) | <0.001* |
| Adjusted | 1.65 (1.30-2.10) | <0.001* |
| **Rural** |  |  |
| Unadjusted | 3.14 (1.80-5.49) | <0.001* |
| Adjusted | 3.39 (1.94-5.93) | <0.001* |
| Adjusted for age, gender, ethnicity, and IMD |  |  |

**Table 10.** Objective 3 Stratified analyses in controls Stratified by IMD.

|  | HR (95% CI) | p-value |
| --- | --- | --- |
| **Most deprived** | 0.98 (0.73 – 1.32) | 0.91 |
| **Least deprived** | 1.60 (1.17 – 2.18) | <0.001* |
| Adjusted for age, gender, ethnicity, and IMD |  |  |

**Figure 1.**
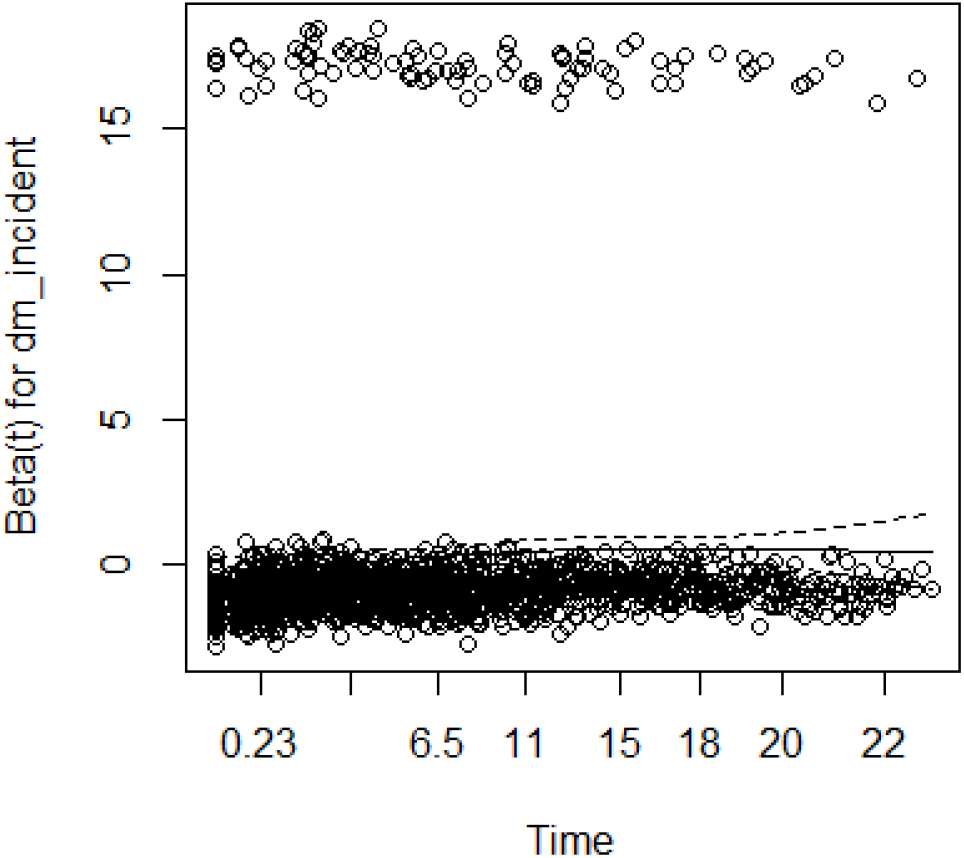
Schoenfeld residual plot

